# A Novel Fixel-Based Approach for Resolving Neonatal White Matter Microstructure from Clinical Diffusion MRI

**DOI:** 10.64898/2026.03.17.26348387

**Authors:** Benjamin T. Newman, Meghan H. Puglia

## Abstract

**Introduction:** Preterm birth is a major risk factor for disrupted brain development and subsequent neurodevelopmental disorders, yet the underlying mechanisms remain poorly understood. Further, typical neuroimaging analyses are particularly challenging in the neonatal brain: data is frequently low quality and a lack of cellular development violates the assumptions relied on by many commonly-used techniques. In this study, we develop and present an advanced diffusion magnetic resonance imaging method to examine the microstructural organization of white matter in a clinically-acquired cohort of premature neonates.

**Methods:** Using a novel approach that resolves multiple tissue compartments within the brain, we provide highly detailed orientation and quantification of white matter fibers and tissue signal fraction. We also utilize a series of automated segmentation algorithms to identify and measure these metrics across key tracts and subcortical regions. We investigate how these measures relate to postmenstrual age, as well as to clinical factors reflecting neonatal illness severity.

**Results:** We report successful segmentation and reconstruction of numerous white matter tracts throughout the neonatal brain. We further demonstrate the utility and functionality of microstructural analysis in a variety of pathologies commonly encountered in the neonatal clinical environment. Our results demonstrate tract-specific developmental trajectories, with early-maturing pathways showing higher microstructural organization. Exploratory analyses suggest that neonatal illness severity has modest, tissue-specific associations with microstructural properties.

**Discussion:** This work demonstrates that advanced microstructural imaging methods can extract meaningful white matter measurements from clinically-acquired scans, providing a practical framework for studying neonatal brain development in real-world hospital settings. These metrics are able to be calculated at extremely young ages, potentially allowing non-invasive study of vulnerable populations before detailed behavioral or neurological assessments are feasible.

## 1 Introduction

Preterm birth, defined as delivery before 37 weeks’ gestation, affects roughly 10% of births in the United States (Centers for Disease Control and Prevention, 2022) and is a leading contributor of long-term neurodevelopmental disorders (Schieve et al., 2016). Infants born preterm are at heightened risk for a broad range of cognitive, motor, and socio-emotional impairments, including learning disabilities, attention-deficit/hyperactivity disorder, and autism spectrum disorder (Johnson, 2007; Lindström et al., 2011; Moreira et al., 2014, 2014; Agrawal et al., 2018; Brydges et al., 2018; Crump et al., 2021). Despite this elevated risk, the mechanisms linking preterm birth to later neurodevelopmental outcomes are not fully understood.

A hallmark of preterm brain injury is disruption to white matter (Ball et al., 2012; Guo et al., 2017), which is particularly vulnerable during the late gestational period when axonal growth, oligodendrocyte maturation, and myelination are actively occurring (Ortinau and Neil, 2015; Bouyssi-Kobar et al., 2016; Volpe, 2019). Diffusion MRI (dMRI) has emerged as a powerful tool for quantifying white matter microstructural features at the subcellular level. This technique measures the movement of water molecules within brain tissue, which is influenced by cellular structures such as axons, myelin, and surrounding extracellular space (Le Bihan and Iima, 2015). From these diffusion patterns, researchers can derive measures that reflect different aspects of tissue organization. For example, fiber density estimates the relative amount of axonal density and fiber diameter within a white matter pathway, providing an indicator of the number or packing of fibers in a tract (Raffelt et al., 2017a). 3-Tissue Constrained Spherical Deconvolution (3T-CSD) signal fractions reflects the proportion of the diffusion signal arising from different tissue compartments, such as intracellular isotropic or gray matter-like tissue, intracellular anisotropic or white matter-like tissue, or extracellular isotropic cerebrospinal fluid-like signal (CSF), offering insight into the relative composition of brain tissue (Frigo et al., 2020). These microstructural compartments have been shown to vary across the lifespan (Newman et al., 2025c)and developmental period (Newman et al., 2023), and to correlate with pathological changes in cellular integrity (Dhollander et al., 2017; Kelly et al., 2022; Newman et al., 2025a). Together, these measures provide complementary information about the structural integrity and organization of neural pathways and can help characterize early brain development.

Prior diffusion studies in preterm neonates have consistently demonstrated delayed maturation and altered connectivity in key white matter tracts. These studies have identified microstructural differences in major projection and association pathways, including the corticospinal tracts, thalamocortical tracts, corpus callosum, and superior longitudinal fasciculi (Ball et al., 2012, 2013; Braga et al., 2015; Guo et al., 2017; Pannek et al., 2018; Pecheva et al., 2019) that have been linked to poorer motor, language, and cognitive outcomes in early childhood (Spittle et al., 2011; Keunen et al., 2017; Salvan et al., 2017). Collectively, these findings suggest that diffusion MRI can detect subtle microstructural disruptions in white matter organization that may precede overt neurological or developmental impairments, offering a window into the early neural substrates of preterm risk.

Despite these prior successes, neonatal brains pose unique challenges for traditional diffusion imaging: myelin is largely undeveloped, extracellular water is abundant, and directional diffusion signals are weak and easily obscured (Dubois et al., 2021). Clinical imaging settings introduce additional limitations, including low b-values and limited angular resolution, which result in lower image detail and fewer angles of measurement, further complicating reliable microstructural estimation (Vos et al., 2016).

To address these challenges in neonatal dMRI analyses (Verschuur et al., 2025), we introduce a novel single-shell three-tissue constrained spherical deconvolution (SS3T-CSD) approach, at low b-values, and using a highly restricted WM response function, to quantify both intra-axonal and extra-axonal diffusion signals in a cohort of preterm neonates. This method allows for refined estimation of fiber orientation distributions, detailed fiber tracking, and calculation of advanced fixel-based metrics of white matter integrity. Using this approach, we examine how white matter microstructure relates to postmenstrual age to better understand early brain maturation following preterm birth, as well as how it relates to clinical factors and neonatal outcomes to identify potential influences on long-term neurodevelopment. We also demonstrate the utility of the method in atypical neuroanatomy, such as neonates with non-standard geometries, and provide a preterm neonatal fiber orientation distribution template to facilitate future stereotaxic alignment and fixel-based analyses.

## 2 Materials and Methods

### 2.1 Participants

Participants were drawn from an ongoing prospective cohort study of infants admitted to the University of Virginia Neonatal Intensive Care Unit (NICU). The study protocol was approved by the University of Virginia Institutional Review Board for Health Sciences Research (HSR210330). Written informed consent was obtained from the parents or legal guardians of all participants prior to participation.

For the present analysis, we included 54 infants from this cohort who underwent clinically-acquired diffusion tensor imaging (DTI) during routine care. Imaging was obtained as part of routine clinical care and not specifically for research purposes. 49 infants passed manual quality control checks for excessive motion or distortion, and a further 6 were removed during the analysis process for poor masking or anatomical segmentation. The final analytic sample consisted of 43 infants.

### 2.2 Diffusion Image Preprocessing

DICOM images were obtained directly from the University of Virginia Health System’s Picture Archiving and Communication System (PACS). Images were anonymized and organized using dcm2niix for nifty conversion and gradient table extraction (Rorden, 2025). Masking and brain extraction was performed directly on the mean b=0 image using a specially adapted version of the Rapid Automatic Tissue Segmentation tool (Oguz et al., 2014). The brain extraction algorithm was initialized using an estimate of the 50^th^ percentile brain volume at birth 340,000 mm^3^ as reported in the literature (Holland et al., 2014), with reductions in initialization volume providing superior results for the most extremely preterm infants. All diffusion images were then denoised (Veraart et al., 2016), Gibbs rings were removed (Kellner et al., 2016), and motion correction with outlier replacement was performed according to established preprocessing protocols (Andersson et al., 2016). No reverse-phase images were collected as part of the clinical process, however *eddy* was utilized for motion correction and could be used with *topup* for distortion correction if the appropriate acquisition was collected (Jenkinson et al., 2012). Images were then uniformly upsampled to a spatial resolution of 1x1x1mm^3^ to minimize partial voluming effects and enforce isotropic voxels to harmonize different clinical acquisitions (Greenspan, 2009; Newman et al., 2020a). This preprocessing step also improves the resolution and interpretability of fiber orientation distributions, enabling clearer identification of crossing fibers (Figure 1).

**Figure 1:**
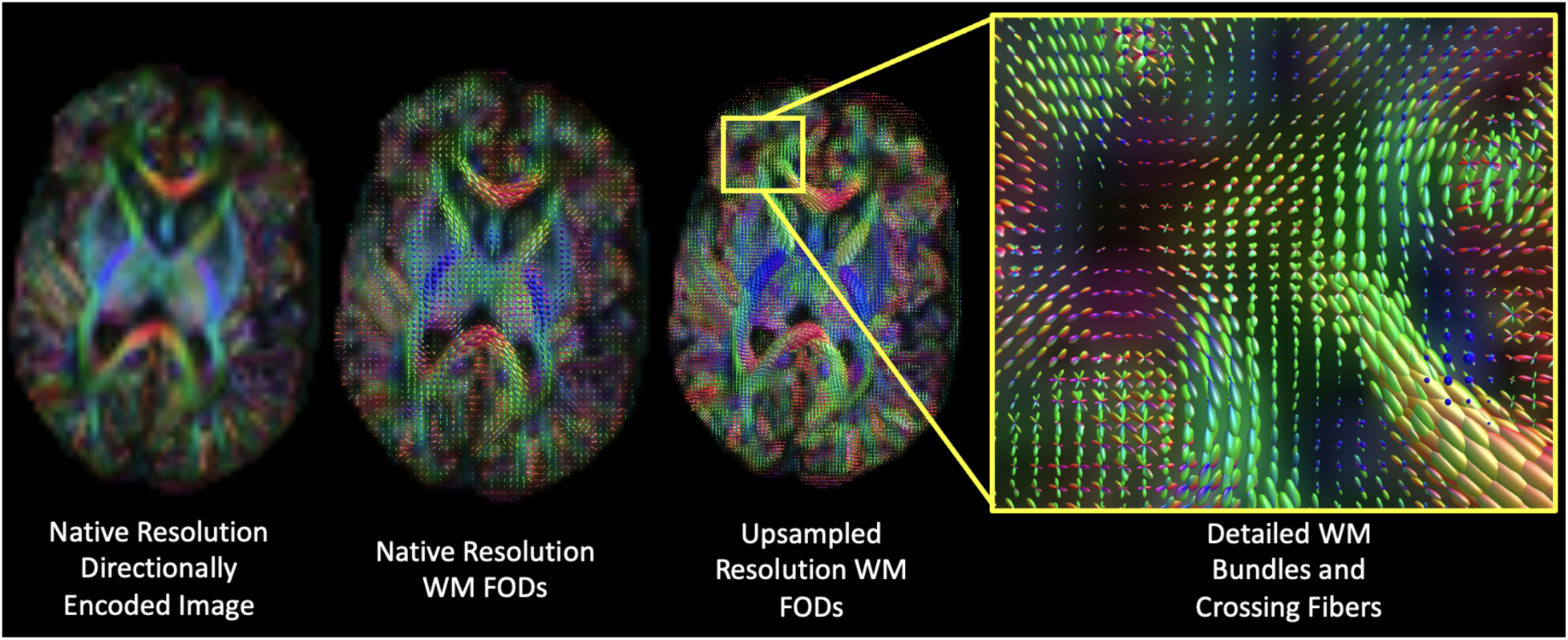
White matter fiber orientation distribution detail improved via upsampling. The preprocessing techniques presented in this study are vital to generating highly detailed white matter FODs and enable the detection of crossing fibers.

#### 2.2.1 Constrained Spherical Deconvolution Application

Following preprocessing, the diffusion images were analyzed using only the b=0 and the b=1000 s/mm^2^ shell. This may initially appear counterintuitive, especially as the developing human connectome project acquired an outer shell of b=3000 s/mm^2^ (Pannek et al., 2012) and when compared to typical adult and adolescent dMRI imaging specifications. It has also been reported that DTI measured fractional anisotropy (FA) values are stable in neonates up to b=3000 s/mm^2^ (Dudink et al., 2008). However, the lower b-value shell provides superior signal-to-noise ratio and ability to resolve poorly myelinated axonal fibers compared to higher b-value shells where the already low directional white matter signal can be overwhelmed by noise. This lower b-value shell does include significantly greater isotropic and extracellular signal, however we have implemented several innovative approaches to identify and remove isotropic signal. This is also a far more typical acquisition for clinical purposes, where speed is prioritized to reduce the time a vulnerable neonate must be physically in the scanner, and thus our results will be more widely applicable to both retrospective and prospective studies.

A further issue that must be confronted in neonatal dMRI, specifically when applying CSD-based models, is the inhomogeneity of white matter fibers across the developing neonatal brain. CSD-based techniques assume that the white matter fiber orientation distribution function can be obtained by deconvolving within each voxel in the brain a single shared common single fiber response function (Tournier et al., 2007; Jeurissen et al., 2014). This response function is typically not defined *a priori,* and is instead derived from a group-wise average value extracted from ‘single fiber voxels,’ which are typically voxels with the highest FA (Dhollander et al., 2016). In the neonatal milieu, the assumptions underlying both CSD and response function definition can be themselves problematic and lead to poor detection and deconvolution of white matter throughout the brain. First, inhomogeneities between white matter fibers are a known issue in adult brains affecting CSD performance (Parker et al., 2013), and neonates display even greater degrees of temporal and spatial white matter inhomogeneities (Bastiani et al., 2019). FA has been found to be far lower throughout the neonatal brain compared to the adult brain (Dudink et al., 2008; Guo et al., 2017; Toselli et al., 2017), which may bias the white matter response function.

Secondly, and a related problem, is that in the absence of well-myelinated axonal fibers, there is a generalized high level of extracellular fluid and isotropic diffusion signal (indicated by increased mean diffusivity) throughout the neonatal white matter skeleton and brain (Bartha et al., 2007; Kimpton et al., 2021). A third issue is that signal from isotropic fluid remains relatively magnified at b=1000 s/mm^2^, with intra-axonal signal having a similar intensity to some extra-axonal signals (Cihangiroglu et al., 2011; Jeurissen et al., 2014). These extra-axonal signals have traditionally been heavily restrictive on fiber tracking and make detection and resolution of crossing fibers difficult (Makki and Hagmann, 2017). While prospective studies in more mature populations can simply acquire higher b-value shells to address this problem, for reasons both clinical, retrospective, and anatomical, an effective methodology for studying the neonatal brain must address these weaknesses at low b-values.

Our approach addresses these issues with several innovations. The workhorse at the center of this approach is single shell 3-tissue constrained spherical deconvolution (SS3T-CSD) (Dhollander and Connelly, 2016a; Dhollander et al., 2018). While other CSD and diffusion models generally perform a single fitting step to describe the diffusion signal, SS3T-CSD instead iterates between two microstructural tissue compartments for three complete cycles. For each iteration, the algorithm fits two response functions to the observed signal, which is always assumed to be isotropic in nature. First, extracellular (CSF-like) signal is separated from intracellular signal by fitting both CSF and grey matter-like response functions to the observed signal. Then, the white matter response function is separated from the remaining intracellular (grey matter-like) signal. This ‘cleans’ the raw diffusion signal with each iteration, identifying directional white matter-like signal and removing isotropic signal that can occlude the relatively weak neonatal white matter signal. A further benefit of the SS3T-CSD approach to both clinical and neonatal data is that the contribution of b=0 images is weighted to 10% of the total signal, rather than an equal weight contributed from all shells as in standard multi-shell multi-tissue CSD (MSMT-CSD) (Jeurissen et al., 2014). This alteration of weights greatly assists in identifying and removing the extracellular CSF-like fluid that is present throughout the neonatal brain, as CSF-like fluid both experiences the highest intensity at b=0 and the greatest signal drop-off as b-values increase. Minimizing the b=0 contribution to total signal thus acts to suppress extracellular isotropic signal, while SS3T-CSD iteratively identifies and separates this signal from intracellular signal, improving white matter resolution.

In order to be effective, however, SS3T-CSD must also correctly identify white matter signal, despite the inhomogeneities across white matter fibers in the neonatal brain. To combat this, we have defined our white matter response function *a priori* using the Dhollander (2019) algorithm. This approach defines an artificial ‘extreme’ single fiber response function using zonal spherical harmonics and further penalizes the presence of extracellular fluid in locating and defining a cohort-specific white matter response function. This approach was demonstrated to significantly improve white matter identification and detection, greatly increasing contrast between white and grey matter in neonates and infants (Dhollander et al., 2019). Response functions for white matter, grey matter, and CSF were defined in each subject in this study, then averaged together to generate mean response functions for use in SS3T-CSD. Both the response function selection algorithm and SS3T-CSD are available from MRtrix3Tissue (https://3tissue.github.io/), a fork of MRtrix3 (Tournier et al., 2019).

#### 2.2.2 Post-processing and Microstructural Analysis

Processing each subject with SS3T-CSD yielded a fiber orientation distribution (FOD) for white matter, grey matter, and CSF. The intensities of these FODs were then log normalized using the *mtnormalise* function in MRtrix3 to correct for residual intensity inhomogeneities (Dhollander et al., 2021) and microstructure signal fraction maps created by summing the coefficients of each white matter, grey matter, and CSF FODs within each voxel to equal 1 (Newman et al., 2020b). Representative output maps from a neonatal subject are shown in Figure 2. The normalized white matter FODs from subjects without severe anatomical deficits were used to generate a group-wise mean population template for preterm neonates using the population_template command. Fixel-based fiber density was calculated for each subject according to established methods (Raffelt et al., 2017b).

**Figure 2:**
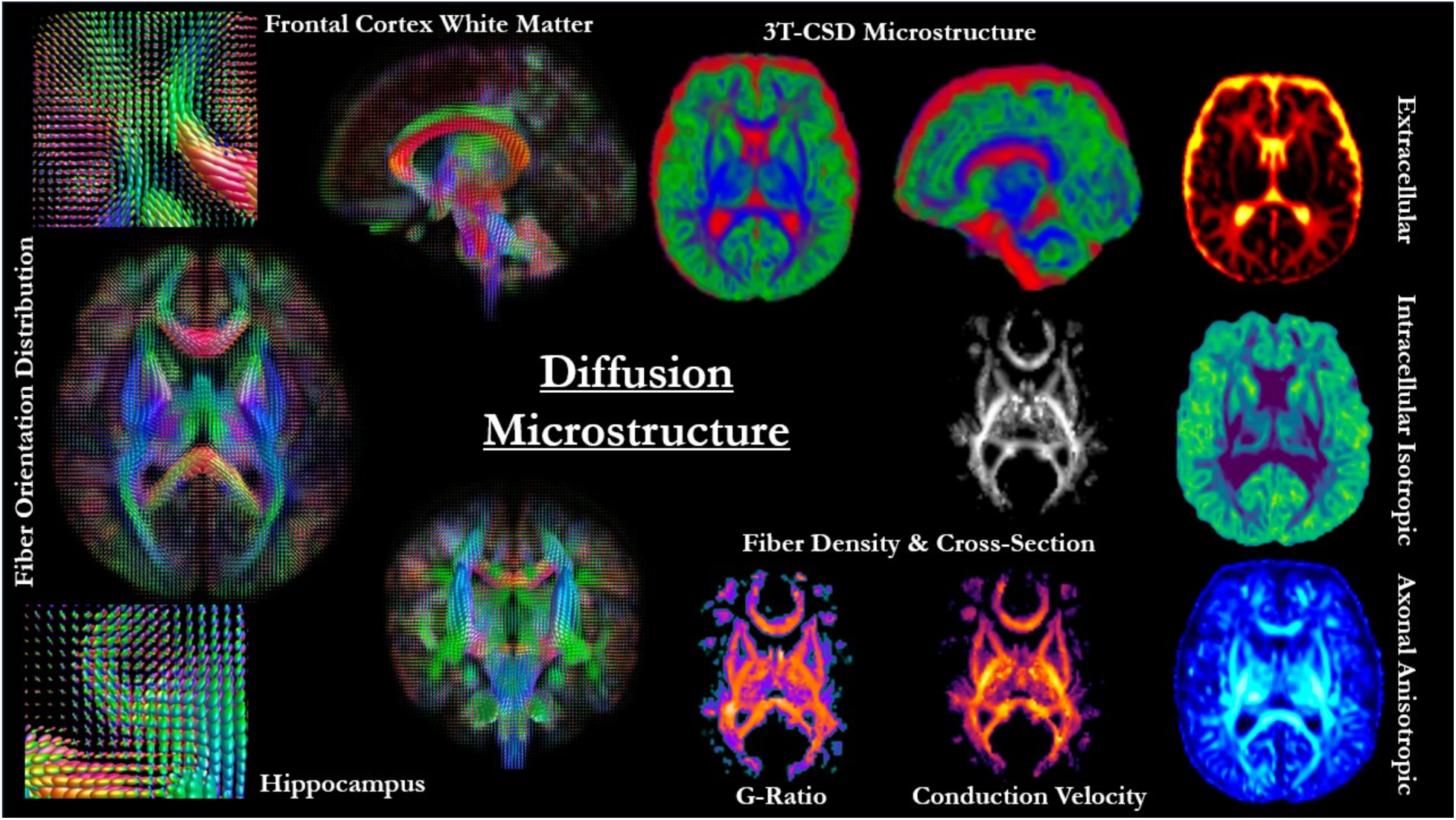
Representative output microstructure maps. Using techniques described in this study, we are able to detect and resolve axonal white matter fibers throughout the brain and generate a wide variety of diffusion microstructure maps from both 3T-CSD and fixel-based techniques. These can potentially be combined with suitable T1- and T2-weighted imaging to calculate g-ratio and axonal conduction velocity maps.

#### 2.2.3 Whole Brain Automatic Segmentation and Tractography

Automatic brain segmentation and parcellation is a priority for objective evaluation of subcortical structures. This process has been considered highly challenging in neonates and infants due to inverted T1w contrast between grey and white matter, with higher extracellular fluid levels in white matter causing these areas to appear darker than grey matter (Zhang et al., 2019). Automated contrast-based methods or even more advanced machine learning based methods trained on older individuals frequently struggle. A recent series of open challenges at the International Conference on Medical Image Computing and Computer Assisted Intervention (MICCAI) has attempted to address this problem in six-month old infants (Wang et al., 2019; Sun et al., 2021). Further issues arise when considering that clinically collected T1w images are frequently differentially sized and positioned compared to dMRI acquisitions, with much wider FOV and resolutions that make co-registration challenging. To address these challenges and in order to employ algorithms and methods with whole lifespan translational benefits, we applied weighted contrast to the diffusion signal fraction maps to generate subject specific images with adult-like T1w contrast (Figure 3). This method has previously been demonstrated to provide similar distributions of intensity to T1w MPRAGE images in healthy adult subjects (Dhollander and Connelly, 2016b). Relative contrast was selected based on experimentally reported peak values for each tissue type, with weights of 95, 60, and 25 for white matter, grey matter, and CSF, respectively based off of previously published results (Song et al., 2026). The resulting image was then segmented into white matter, cortical grey matter, and subcortical grey matter using FSL’s *FIRST* and *FAST* hidden Markov, Bayesian, and expectation-maximization algorithms including the average b=0 image as T2-weighted contrast and implemented in the *5ttgen* MRtrix3 function (Zhang et al., 2002; Smith et al., 2004, 2012; Patenaude et al., 2011). Individual subcortical structures (Figure 4) were obtained from FIRST and FAST surface segmentations.

**Figure 3:**
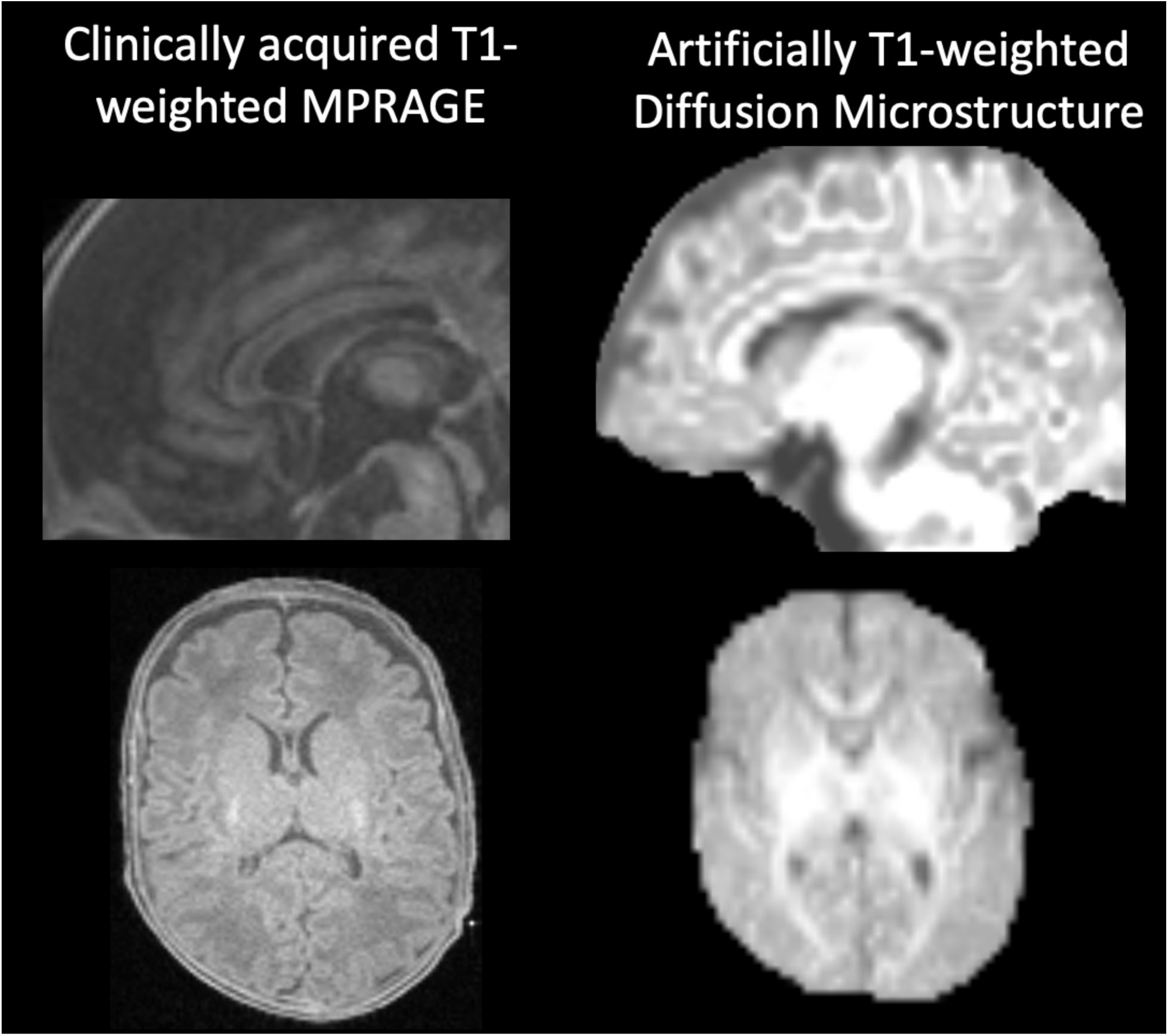
Comparison of clinically acquired T1-weighted MPRAGE and artificial contrast corrected diffusion microstructure map. Artificial maps with adult-like T1-weighted contrast were generated by re-weighing CSF, white matter, and grey matter signal fraction maps to facilitate the use of established intensity-based segmentation techniques.

**Figure 4:**
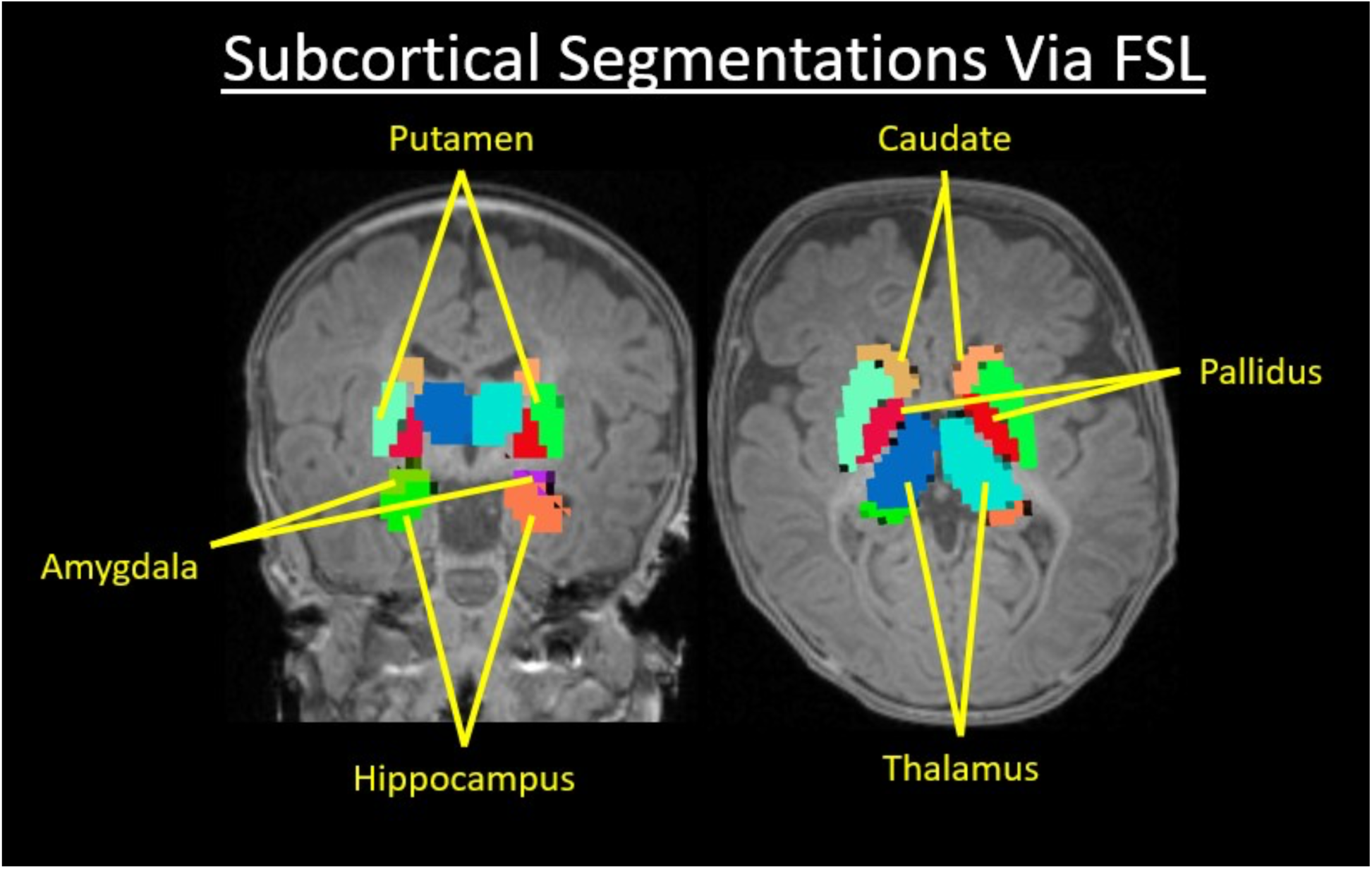
Subcortical structures segmented from artificially generated T1-weighted contrast using 3T-CSD diffusion microstructure maps. Using FSL’s FIRST and FAST automated segmentation algorithms, 7 bilateral subcortical structures (including the Nucleus Accumbens, not labeled above) were able to be accurately segmented across participants.

The resulting 5 Tissue Type (5tt) image was then used to perform Anatomically Constrained Tractography (ACT) using the probabilistic iFOD2 algorithm to dynamically seed 10,000,000 tracts. The 5tt image when used in ACT constrains tracts to voxels identified as white matter with minimum length of 2mm and maximum curvature of 45°. Spherical informed filtering of tractograms (SIFT) was used to reduce the number of tracts to 1,000,000 based on underlying white matter-FOD lobe integrals (Smith et al., 2013). Additionally, for more accurate segmentation of specific white matter tracts, each subject’s white matter FODs were converted to spherical harmonic peaks using the *sh2peak* command in MRtrix3 (Jeurissen et al., 2013) in order to be used in the TractSeg convolutional neural network-based segmentation approach (Wasserthal et al., 2018). TractSeg was implemented on the GPU architecture maintained by University of Virginia Research Computing and used the most recently-released set of training weights to segment each subject’s white matter into up to 72 tract bundles (Figure 5). Fixel-based fiber density, and 3T-CSD white matter-like, grey matter-like, and CSF-like microstructure signal fractions were averaged across all voxels in each whole brain region of interest (ROI), subcortical grey matter region, and tract bundle in each subject for statistical analysis.

**Figure 5:**
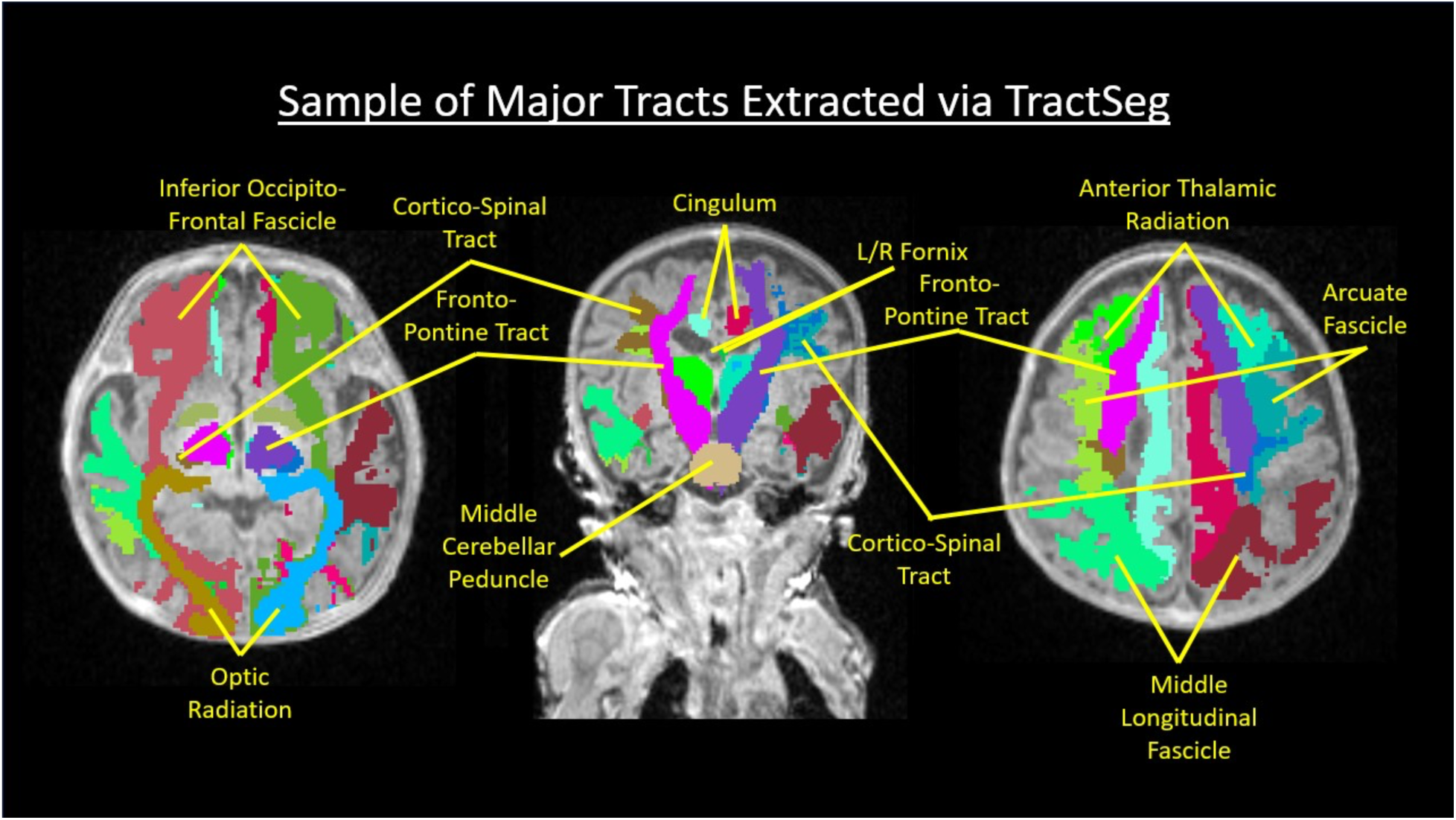
Major white matter tracts identified and extracted from neonatal FODs using the TractSeg neural network. Resolution of white matter FODs is sensitive and detailed to define and extract richly detailed maps of major white matter tracts and fasciculi.

### 2.3 Statistical Analysis

Planned analyses were preregistered (aspredicted.org, #276926) prior to data inspection. All statistical analyses were conducted in R version 4.3.1 (R Core Team, 2020) using the lme4 (Bates et al., 2015) and lmerTest (Kuznetsova et al., 2017) packages to assess fiber density in white matter tracts and tissue signal fraction in subcortical structures.

Tracts and structures with >50% zero values across subjects were excluded prior to analysis. These included corpus callosum regions and commissures (anterior commissure, rostrum, rostral body, anterior midbody), the fornix, inferior cerebellar peduncle, striatal–cortical tracts (striato-premotor, striato-fronto-orbital, striato-precentral, striato-postcentral), superior longitudinal fascicles (I–III), thalamo-premotor and thalamo-postcentral tracts, the superior thalamic radiation, and the uncinate fascicle.

Remaining tracts were classified according to established developmental trajectories (Ball et al., 2013; Braga et al., 2015). Early-maturing tracts included cerebellar peduncles, corticospinal pathways, thalamo-cortical tracts, optic radiation, posterior corpus callosum regions, and pontine projections. Later-developing association tracts included superior, middle, and inferior longitudinal fascicles, the arcuate fascicle, inferior occipito-frontal fascicle, cingulum, striatal–cortical projections, anterior thalamic radiation, and the genu of the corpus callosum.

Subjects with zero values in >25% of the remaining tracts (n = 6) or subcortical structures (n = 3) were excluded. Outlier values (>3 median absolute deviations above the median) were also removed, accounting for 13.3% of tract data points and 9.6% of subcortical structure data points.

Linear mixed-effects models were fit using restricted maximum likelihood (REML), with significance of fixed effects assessed via Satterthwaite’s approximations for degrees of freedom. Model diagnostics, including residual distributions, random-effects structure, and singularity checks were performed to ensure model appropriateness.

All models included random effects for subjects to account for repeated measurements. For white matter tract fiber density, an additional random effect allowed each tract to vary within subject, as ANOVA comparisons confirmed that including tract-level variability significantly improved model fit. For tissue signal fraction in subcortical structures, only subject-level random effects were included because structure-level random effects exhibited low variance and resulted in singular model fits. Residual inspection further indicated that zero values distorted model estimation; therefore, remaining zero values were excluded prior to model fitting. Where possible, the same random-effects structure was used across analyses for comparability, with deviations due to singularity noted. All models included mean-centered postmenstrual age at MRI (days), mean-centered gestational age at birth (days), and sex as covariates.

#### 2.3.1 Description of white matter microstructure

Descriptive statistics (mean and standard deviation) were computed for fiber density by tract and tissue signal fraction by subcortical structure to characterize the distribution, central tendency, and variability of tract-specific fiber density and tissue fraction metrics across the cohort. As a methodological validation step, we evaluated whether fiber density values reflected established neonatal white matter maturation patterns by comparing early-maturing projection and commissural tracts with later-developing association tracts. Mean fiber density values were compared between these tract classes using Welch’s two-sample t-test. Tissue signal fractions were evaluated using a repeated-measures ANOVA with tissue type (CSF, grey matter, white matter) as a within-subject factor, followed by Bonferroni-corrected paired comparisons to assess expected tissue-specific distributions.

#### 2.3.2 Maturation of white matter microstructure

To test the hypothesis that fiber density in early-maturing tracts would increase with postmenstrual age and later-developing association tracts would show lower fiber density at comparable postmenstrual age, we included the interaction between postmenstrual age and tract classification (early vs. late) as a fixed effect in a linear mixed-effects model. Because eight subjects had postmenstrual age values of approximately one year or greater (range 363– 1459 days), a complementary model was performed in a restricted sample of younger participants (postmenstrual age < 360 days, range: 239–307) to ensure that these relative outliers did not drive observed effects.

#### 2.3.3 Maturation of tissue fractions

To test the hypothesis that subcortical grey matter fraction increases and CSF fraction decreases with postmenstrual age, we included the interaction between postmenstrual age and tissue type (CSF, grey matter, white matter) as a fixed effect in a linear mixed-effects model. Three subjects had postmenstrual age values of approximately one year or greater (range 363–370 days). As such, a complementary model was restricted to younger participants (postmenstrual age < 360 days, range: 253–303) to ensure these relative outliers did not drive results.

#### 2.3.4 Exploratory clinical correlates

In exploratory analyses, we examined whether greater illness severity was associated with lower fiber density and higher CSF fraction. Clinical data were extracted from the University of Virginia Health System electronic health record via the Caboodle enterprise data warehouse (Epic Systems, Madison, WI). Illness severity was characterized using the Heart Rate Observation (HeRO) (Fairchild and Aschner, 2012), Neonatal Sequential Organ Failure Assessment (nSOFA) (Berka et al., 2022), and Prognostic Respiratory Intensity Scoring Metric (PRISM) (Nance et al., 2025) scores.

For HeRO, three metrics were considered: the value closest to the MRI date, the value closest to birth, and the maximum score recorded during hospitalization as an index of peak illness burden. For nSOFA, single time-point values showed minimal variability across participants (97.37% zero values); therefore, only maximum nSOFA scores were analyzed. PRISM scores reflect cumulative illness severity during hospitalization and were therefore analyzed as single summary values.

Our preregistration included the hypothesis that lower fiber density in motor tracts would be associated with delayed gross motor outcomes. However, charted motor outcome measures were sparse in the electronic health record, and as such, this hypothesis could not be evaluated with the current dataset.

Associations between illness severity and imaging measures were tested using linear mixed-effects models including illness severity scores as fixed effects. Additional models examined interactions between illness severity and tract classification for fiber density analyses, and between illness severity and tissue type for tissue signal fraction analyses.

## 3 Results

### 3.1 Participant Characteristics

The analytic sample included 43 neonates (27 female). The majority of participants were White (67%), with 16% identifying as Black or African American, and 16% as Other, multiracial, or Hispanic/Latino. Most infants were singleton births (65%), with 28% twins and 7% triplets. Mean gestational age (GA) at birth was 201.8 ± 31.0 days (∼28.8 ± 4.4 weeks). The majority of participants (53.49%) were classified as Extremely Preterm (< 28 weeks’ GA), with 23.26% born Very Preterm (28–32 weeks’ GA), 16.28% born Moderate to Late Preterm (32–37 weeks’ GA), and 6.98% born Early Term (37–40 weeks’ GA). Mean postmenstrual age at MRI was 282.6 ± 30.7 days (∼40.4 ± 4.4 weeks). Delivery was predominantly by Cesarean section (56%), with 16% born vaginally; delivery method was not reported for the remaining participants. A summary of clinical MRI findings is provided in Table 2.

### 3.2 White matter microstructure derived from clinical-quality scans reproduces expected anatomical and developmental patterns

Our diffusion modeling pipeline produced anatomically coherent whole-brain tractograms from clinically acquired diffusion data. Probabilistic tractography generated dense white matter reconstructions across the neonatal brain, including successful detection and tracking of crossing fibers (Figure 6). Segmentation of these tractograms using TractSeg further enabled identification of major projection and association pathways, including corticospinal tracts, longitudinal fasciculi, and the cingulum (Figure 7), demonstrating that clinically-acquired diffusion scans can support tract-specific analyses using this framework. In addition to enabling tractography and segmentation, the resulting 3T-CSD microstructure maps provided interpretable tissue-specific signatures that could reflect common neonatal neuropathology (Figure 8), illustrating the potential clinical utility of this approach for characterizing brain microstructure in vulnerable populations.

**Figure 6:**
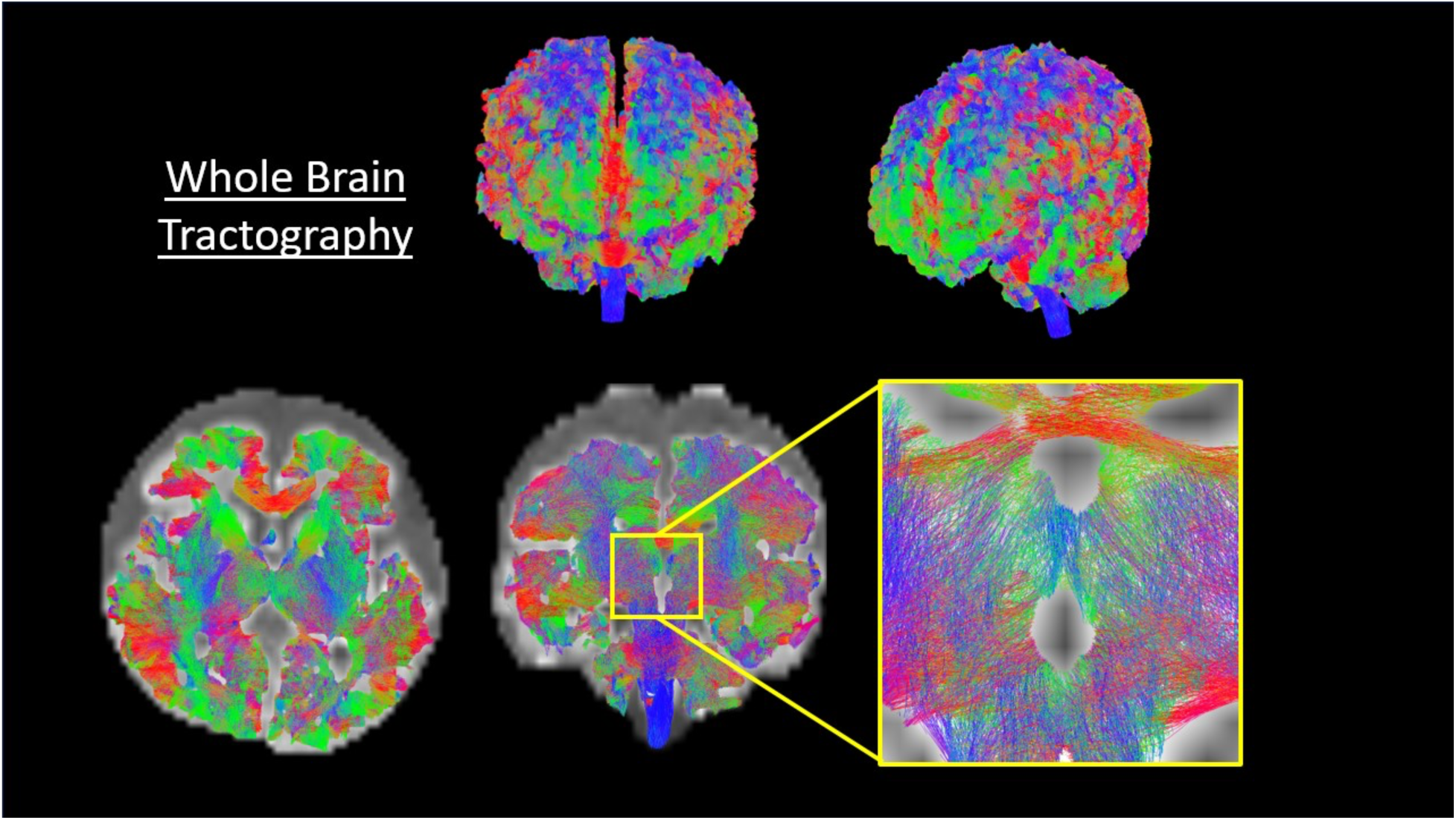
Representative whole brain tractogram. Using probabilistic fiber tracting and established algorithms ACT, iFOD2, and SIFT we were able to generate tracts with similar sensitivity, coverage, and complexity to adult subjects. Crossing fibers were successfully detected and tracked (see insert) while using SIFT permitted evaluation of tracts using underlying axonal anatomical constraints. Resulting tractograms contained 1 million tracts for each participant, permitting connectome-based strength and connectivity comparisons.

**Figure 7:**
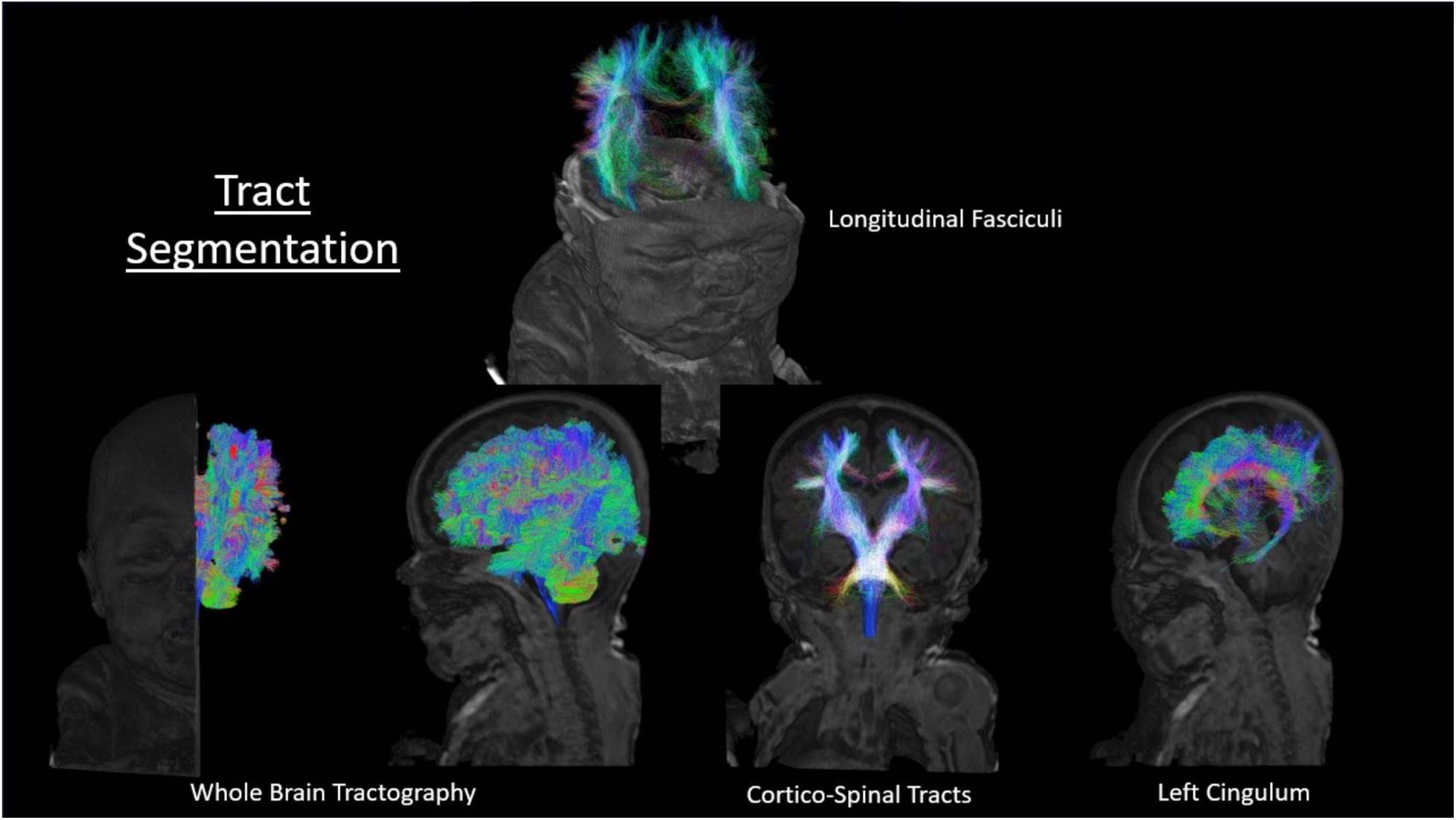
Extraction of major white matter tracts, as extracted via TractSeg, in representative data from the cohort. Combining whole-brain tractograms and TractSeg extracted voxels permits the illustration and quantification of major white matter structures, including longitudinal fasciculi, cortico-spinal tracts, and the cingulum. In addition to demonstrating anatomical accuracy for the entire pipeline, this work potentially opens the door to tract-based spatial statistics and tract-specific measures in the neonatal population.

**Figure 8:**
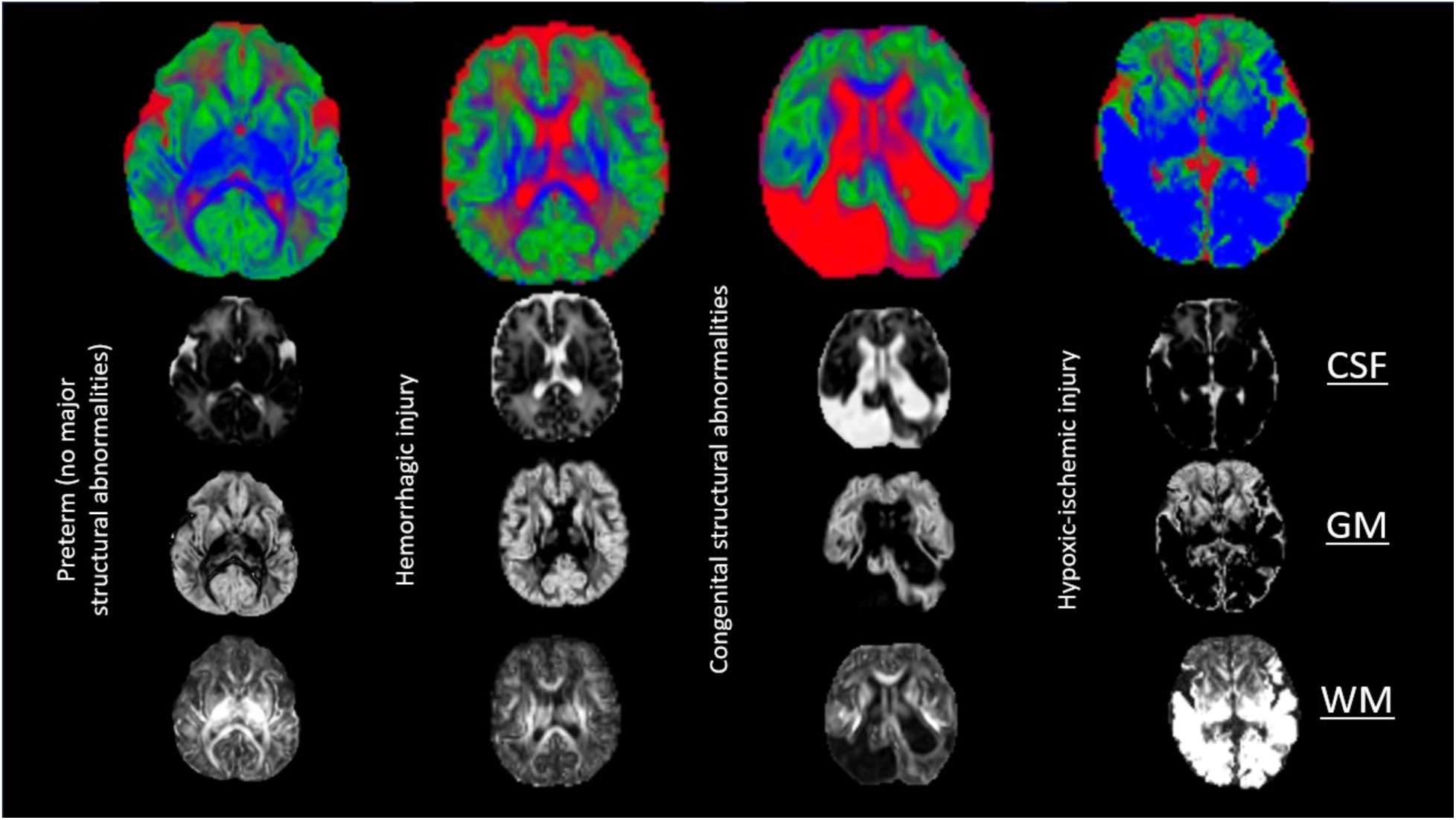
Illustration of various pathologies within the neonatal population and their representation by 3T-CSD microstructural metrics. Illustrative examples of microstructural signal patterns observed across the cohort, including cases with hemorrhage, ventricular enlargement, and ischemic injury.

Summary statistics for fiber density by tract and tissue signal fraction by subcortical structure are provided in Table 1. Fiber density differed between tract classes in the expected pattern. Early-maturing tracts showed higher mean fiber density (0.657 ± 0.252) than later-developing association tracts (0.447 ± 0.160), consistent with neonatal white matter maturation patterns. A Welch two-sample t-test confirmed this difference was highly significant (*t*(1373) = 19.97, *p* < 0.001, 95% CI [0.190, 0.231]).

**Table 1.**
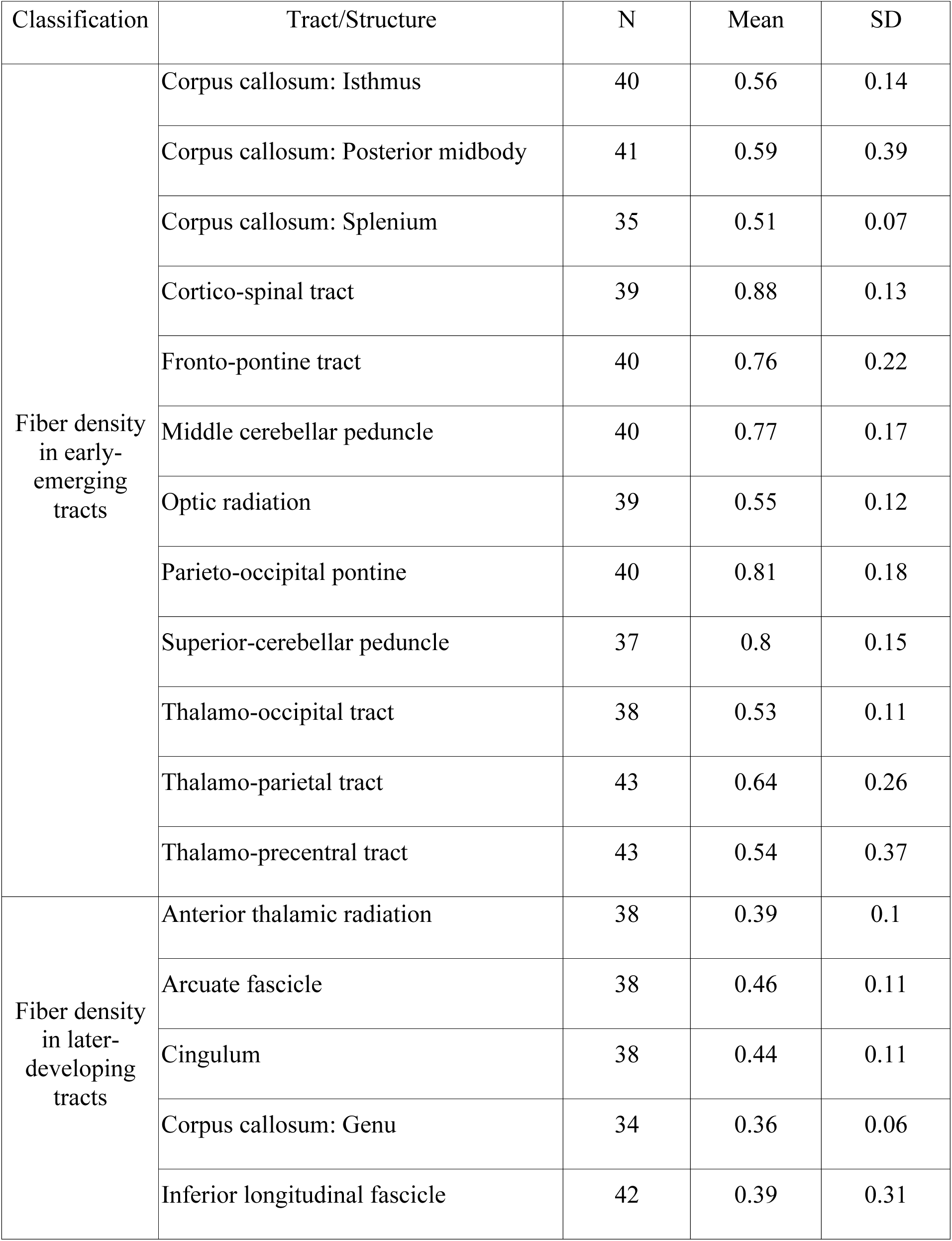
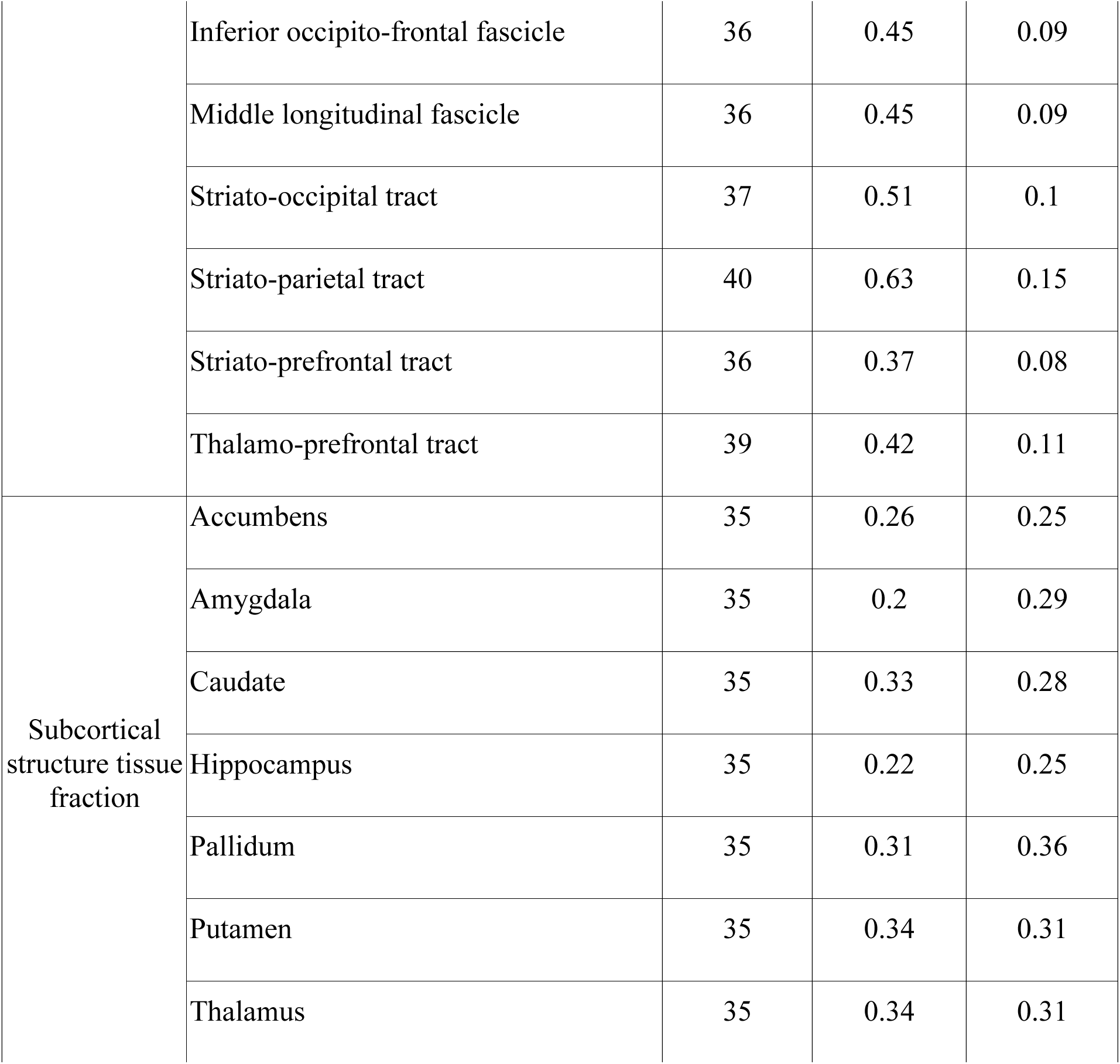
Fiber density and tissue signal fraction summary statistics.

| Classification | Tract/Structure | N | Mean | SD |
| --- | --- | --- | --- | --- |
| Fiber density<br>in early-emerging<br>tracts | Corpus callosum: Isthmus | 40 | 0.56 | 0.14 |
|  | Corpus callosum: Posterior midbody | 41 | 0.59 | 0.39 |
|  | Corpus callosum: Splenium | 35 | 0.51 | 0.07 |
|  | Cortico-spinal tract | 39 | 0.88 | 0.13 |
|  | Fronto-pontine tract | 40 | 0.76 | 0.22 |
|  | Middle cerebellar peduncle | 40 | 0.77 | 0.17 |
|  | Optic radiation | 39 | 0.55 | 0.12 |
|  | Parieto-occipital pontine | 40 | 0.81 | 0.18 |
|  | Superior-cerebellar peduncle | 37 | 0.8 | 0.15 |
|  | Thalamo-occipital tract | 38 | 0.53 | 0.11 |
|  | Thalamo-parietal tract | 43 | 0.64 | 0.26 |
|  | Thalamo-precentral tract | 43 | 0.54 | 0.37 |
| Fiber density<br>in later-developing<br>tracts | Anterior thalamic radiation | 38 | 0.39 | 0.1 |
|  | Arcuate fascicle | 38 | 0.46 | 0.11 |
|  | Cingulum | 38 | 0.44 | 0.11 |
|  | Corpus callosum: Genu | 34 | 0.36 | 0.06 |
|  | Inferior longitudinal fascicle | 42 | 0.39 | 0.31 |
|  | Inferior occipito-frontal fascicle | 36 | 0.45 | 0.09 |
|  | Middle longitudinal fascicle | 36 | 0.45 | 0.09 |
|  | Striato-occipital tract | 37 | 0.51 | 0.1 |
|  | Striato-parietal tract | 40 | 0.63 | 0.15 |
|  | Striato-prefrontal tract | 36 | 0.37 | 0.08 |
|  | Thalamo-prefrontal tract | 39 | 0.42 | 0.11 |
| Subcortical<br>structure tissue<br>fraction | Accumbens | 35 | 0.26 | 0.25 |
|  | Amygdala | 35 | 0.2 | 0.29 |
|  | Caudate | 35 | 0.33 | 0.28 |
|  | Hippocampus | 35 | 0.22 | 0.25 |
|  | Pallidum | 35 | 0.31 | 0.36 |
|  | Putamen | 35 | 0.34 | 0.31 |
|  | Thalamus | 35 | 0.34 | 0.31 |

**Table 2.** Summary of clinical MRI findings.

| Category | N | % |
| --- | --- | --- |
| Normal MRI / no intracranial abnormality | 18 | 41.9% |
| Prior hemorrhage / germinal matrix hemorrhage / microhemorrhage | 11 | 25.6% |
| White matter/hypoxic-ischemic injury | 4 | 9.3% |
| Prominent extra-axial or subarachnoid spaces | 6 | 14.0% |
| Ventriculomegaly / hydrocephalus | 3 | 7.0% |
| Major congenital brain malformation | 3 | 7.0% |
| Reduced brain volume / diffuse volume loss | 3 | 7.0% |
| Subdural hemorrhage | 3 | 7.0% |

Subcortical tissue signal fractions also showed the expected distribution across tissue types. A repeated-measures ANOVA revealed a significant effect of tissue type on mean signal fraction (*F*(2,102) = 139.12, *p* < 0.001, generalized η² = 0.732). Bonferroni-corrected paired comparisons indicated that white matter fractions (*M*=0.530) were higher than grey matter (*M*=0.291) and CSF (*M*=0.039), and grey matter fractions were higher than CSF (all *p*s < 0.001).

### 3.3 Developmental increases in fiber density reveal tract-specific maturation patterns

We used a linear mixed-effects model to examine associations between postmenstrual age and tract class on fiber density. Fiber density increased significantly with postmenstrual age (β = 0.0018, SE = 0.00044, *p* < .001) and with GA (β = 0.0014, SE = 0.00040, *p* = .001). Early-maturing tracts showed significantly higher fiber density than later-developing association tracts (β = −0.223, SE = 0.0095, *p* < .001). Sex was not associated with fiber density (*p* = .777).

There was also a significant interaction between postmenstrual age and tract class (β = 0.0010, SE = 0.0003, *p* = .004), indicating that the association between postmenstrual age and signal fraction differed between tract classes. Specifically, later-developing tracts showed a slightly steeper increase in fiber density with age compared to early-maturing tracts (Figure 9).

**Figure 9.**
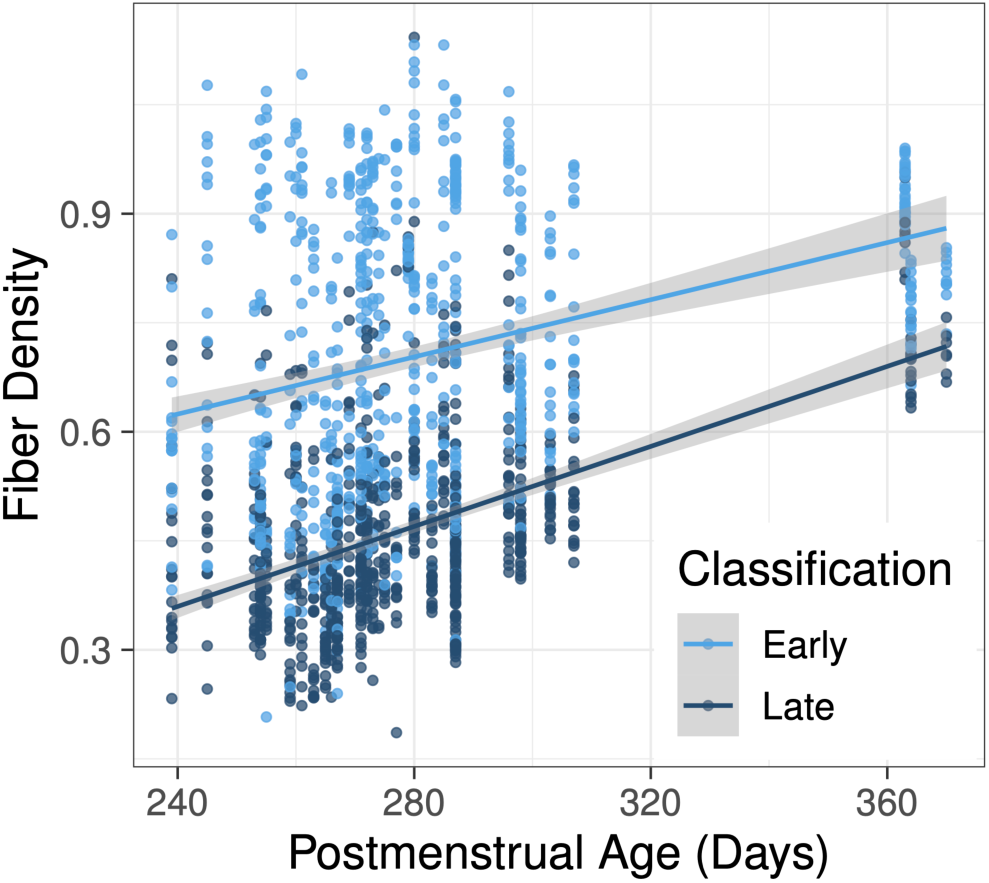
Interaction between postmenstrual age and tract class on fiber density. Fiber density was higher in early-emerging tracts, and increased with postmenstrual age in both early-maturing and later-developing white matter tracts at different rates. Specifically, later-developing association tracts exhibited a slightly steeper increase in fiber density with age compared to early-maturing tracts.

To evaluate robustness of these findings in the younger subset of infants, a parallel model showed similar developmental effects (Supplemental Figure S1). Fiber density increased with postmenstrual age (β = 0.0028, SE = 0.00089, *p* = .003) and GA (β = 0.0014, SE = 0.00045, *p* = .004), and was higher in early-maturing tracts (β = −0.236, SE = 0.011, *p* < .001). However, the interaction between postmenstrual age and tract class did not reach statistical significance in this age-restricted subset (*p* = .654). Sex was again not associated with fiber density (*p* = .583).

### 3.4 Tissue signal fractions differ across brain compositions and show distinct developmental trajectories in white and gray matter

We used a linear mixed-effects model to examine associations between postmenstrual age and tissue type on signal fraction. Signal fraction differed significantly across tissue classes (Figure 10). Relative to CSF, signal fraction was substantially higher in grey matter (β = 0.296, SE = 0.0125, *p* < .001) and white matter (β = 0.569, SE = 0.0125, *p* < .001). Although there was no overall main effect of postmenstrual age (*p* = .395), the association between postmenstrual age and signal fraction differed by tissue type. Specifically, signal fraction increased with postmenstrual age in white matter (β = 0.0045, SE = 0.00042, *p* < .001) but decreased in grey matter (β = −0.0026, SE = 0.00042, *p* < .001). GA (*p* = .887) and sex (*p* = .867) were not significantly associated with signal fraction. 3T-CSD maps illustrating age-related changes in brain composition can be seen in Figure 11.

**Figure 10.**
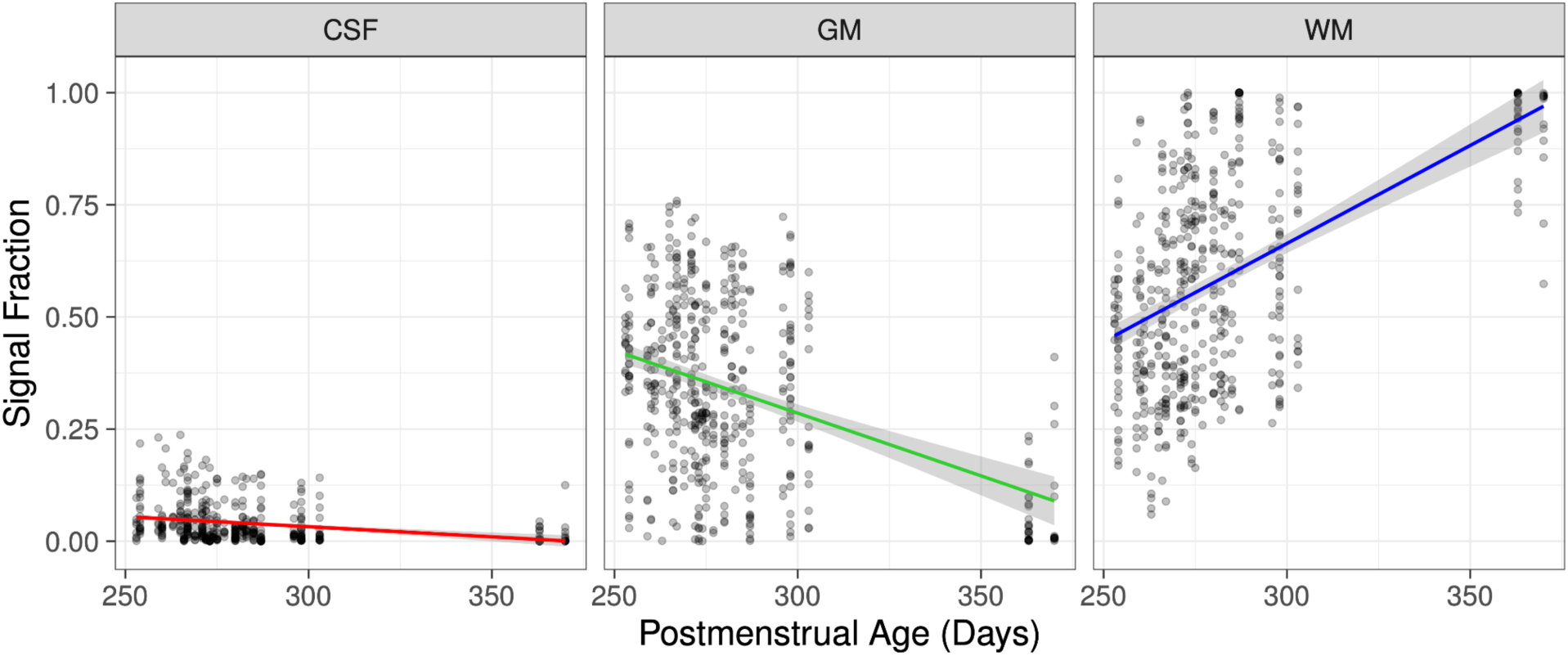
Developmental trajectories of signal fraction differed by tissue type. Signal fraction increased with postmenstrual age in white matter (WM), decreased in grey matter (GM), and demonstrated slight, albeit non-significant, decreases in cerebrospinal fluid (CSF), indicating distinct maturation patterns across brain tissues. Red=CSF-like, green=grey matter-like, and blue=white matter-like signal fraction.

**Figure 11.**
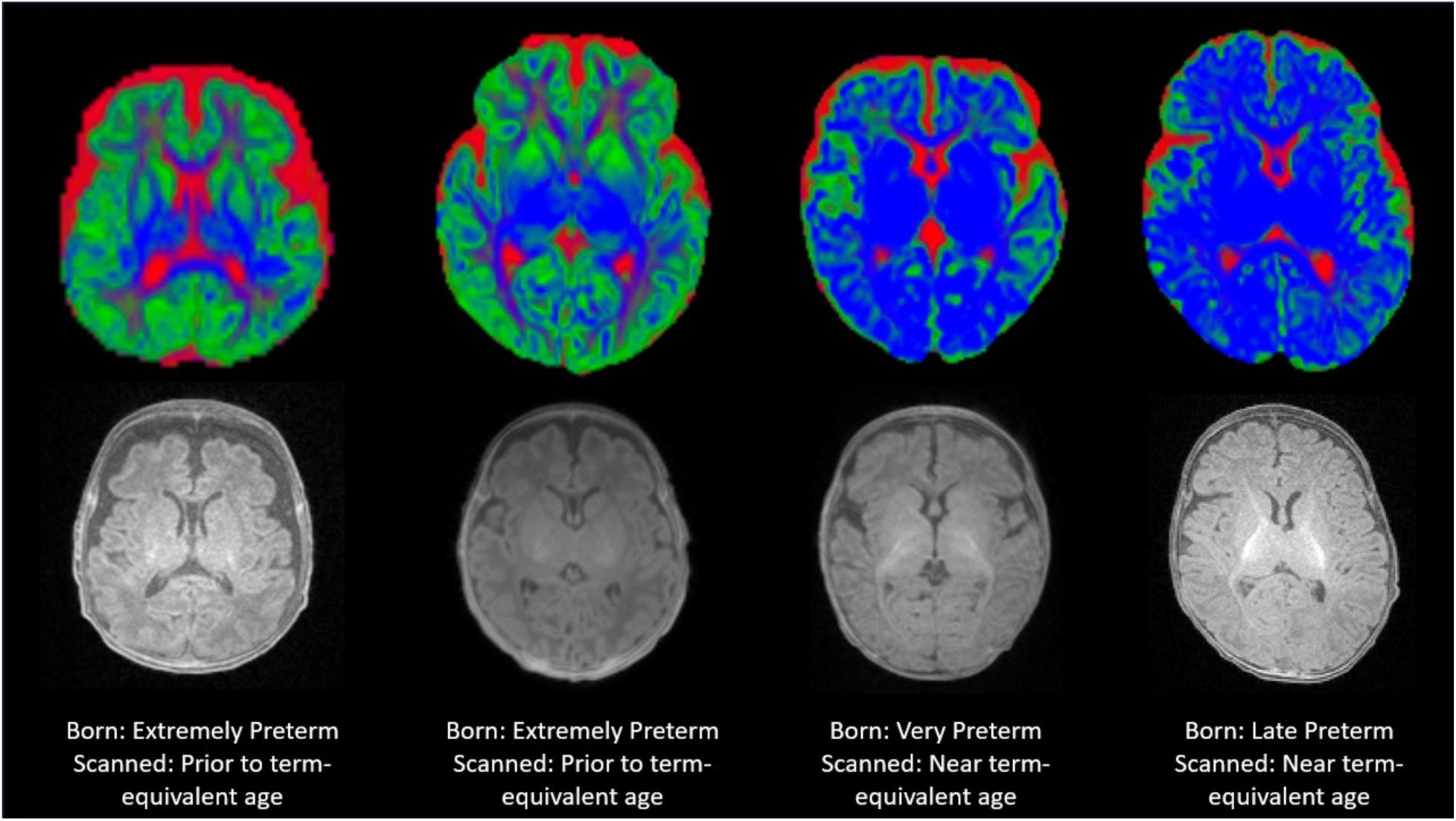
Comparison of age-related changes in brain composition of 3T-CSD maps. Top row: red=CSF-like, green=grey matter-like, and blue=white matter-like; Bottom row: T1-weighted MPRAGE images. Despite relatively similar T1-weighted images and normal clinical reports, the degree of change in 3T-CSD metrics is apparent, with a general increase in white matter-like signal fraction and a decrease in grey matter-like signal fraction across the entire cortex and subcortical grey matter. Development of the frontal cortex is also highly apparent.

Analyses restricted to the younger subset yielded a similar pattern (Supplemental Figure S2). Signal fraction again differed significantly across tissue classes (both *p* < .001). The interaction between postmenstrual age and white matter remained significant (β = 0.0063, SE = 0.0010, *p* < .001), indicating increasing signal fraction with age in white matter. In contrast, the postmenstrual age-by–grey matter interaction was not significant in this subset (*p* = .258). Neither GA (*p* = .657) nor sex (*p* = .714) was associated with signal fraction.

### 3.5 Illness severity shows minimal associations with tissue signal fractions

Exploratory analyses examined whether illness severity was associated with fiber density and tissue signal fraction. Across models including HeRO scores (closest to MRI, at birth, and maximum during hospitalization), nSOFA maximum scores, and PRISM scores, no significant main effects of illness severity were observed for signal fraction after controlling for postmenstrual age, GA at birth, and sex (all *p*s > .20). Illness severity measures also did not interact with tract classification in fiber density models (all *p*s > .28).

In tissue signal fraction models, a significant interaction between HeRO score at birth and tissue type was observed, indicating differential associations across tissues (Figure 12). Specifically, higher HeRO scores at birth were associated with higher grey matter signal fraction (β = 0.0265, SE = 0.0084, *p* = .002) and lower white matter signal fraction (β = −0.047, SE = 0.0083, *p* < .001). In contrast, HeRO scores closest to MRI showed a significant association only with white matter signal fraction (β = −0.103, SE = 0.0354, *p* = .004). Interactions between tissue type and maximum HeRO scores, nSOFA, and PRISM were not significant (all interaction *p*s > .074).

**Figure 12.**
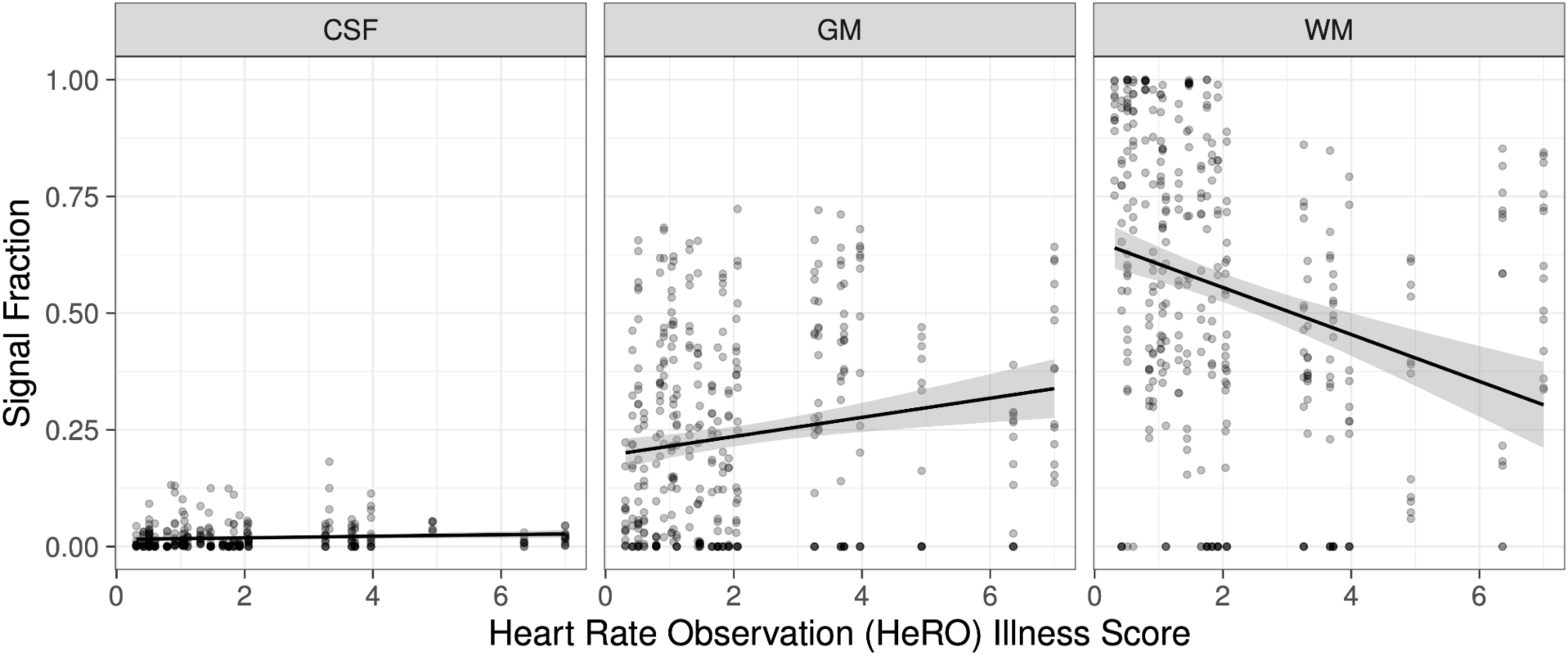
Associations between illness severity and tissue signal fraction. A significant interaction between Heart Rate Observation (HeRO) illness score at birth and tissue type indicated differential relationships across brain tissues. Higher HeRO scores at birth were associated with increased grey matter (GM) signal fraction and decreased white matter signal fraction (WM). There was no association with cerebral spinal fluid (CSF) signal fraction.

## 4 Discussion

This study demonstrates that fixel-based measures of white matter microstructure can be reliably derived from clinically-acquired neonatal diffusion MRI. Using a constrained spherical deconvolution framework adapted for low-resolution clinical scans, we generated fiber density and tissue signal fraction maps that supported tract-specific analyses and subcortical tissue segmentation across the neonatal brain. These measures reproduced expected patterns of early white matter maturation, including higher fiber density in early-developing projection tracts and developmental increases in white matter signal fraction with postmenstrual age. Together, these findings show that clinically-acquired diffusion MRI, even when acquired with lower spatial and angular resolution, can support biologically meaningful microstructural analyses, expanding the potential for advanced diffusion modeling in neonatal and other clinical populations.

Consistent with prior studies of neonatal brain development (Ball et al., 2013; Braga et al., 2015), we observe clear tract-specific maturation patterns. Fiber density was higher in early-maturing projection tracts than in later-developing association pathways, and tissue signal fraction increased with postmenstrual age in white matter while gray matter showed distinct developmental trajectories. These findings indicate that biologically meaningful developmental signals can be detected even in clinically acquired diffusion scans of preterm infants.

Exploratory analyses examined associations between clinical illness severity and microstructural measures. Notably, at birth, higher HeRO scores, a measure derived from continuous heart rate monitoring that indexes autonomic instability and risk for neonatal sepsis, were associated with higher gray matter fraction and lower white matter fraction. In contrast, maximum HeRO scores during hospitalization, as well as nSOFA and PRISM scores, did not show significant associations. The finding that HeRO at birth was the most informative metric suggests that early physiological instability may exert a cascading influence on brain development, highlighting the importance of early insults in shaping subsequent microstructural trajectories (Mathur et al., 2010). The absence of associations with later or cumulative illness severity measures may reflect the sensitivity of HeRO to immediate neonatal stressors that are not captured by broader composite scores. Additionally the selected cohort was recruited exclusively for preterm birth and available imaging; a more specifically-recruited population without the wide variety of complications from preterm birth may be more closely correlated to specific measures.

A key contribution of this work is the demonstration that clinically-acquired MRI scans, which often have lower spatial and angular resolution than research-grade acquisitions, can nevertheless yield meaningful microstructural information when analyzed with advanced multi-compartment modeling techniques. The approach described in this study does not require any specialized acquisitions, relies entirely on publicly-available software, and does not even necessitate the co-acquisition of a T1-weighted image. The resulting tractography, tract segmentation, and microstructural metrics successfully identified and segmented major WM tracts and reproduced established patterns of neonatal WM organization and maturation. These findings suggest that large archives of clinically acquired neonatal diffusion scans may represent an underutilized resource for studying early brain development and the effects of perinatal illness. This approach therefore increases the feasibility of studying vulnerable neonatal populations without requiring research-dedicated imaging sessions, opens the possibility of leveraging large retrospective clinical imaging datasets to investigate early brain development and identify neuroimaging markers of neonatal risk, and provides a practical framework for broader implementation in hospital settings.

In previously published studies, our group has highlighted the ability of diffusion microstructure to detect functionally significant differences in autism spectrum disorder (ASD) (Newman et al., 2024, 2025b; Ressa et al., 2024; Coleman CR, Nance MG, Jacokes Z, Druzgal TJ, Arutiunian V, Kresse A, et al., 2025). Like several other childhood behavioral disorders, ASD is typically diagnosed after behavioral differences manifest in early childhood. Calculation of microstructure metrics in adolescents who go on to receive ASD diagnoses may identify early signs or patterns that improve detection and allow for earlier intervention. This same approach could be used to benefit attention deficit hyper-activity disorder (ADHD), and other early manifesting child behavioral disorders.

### 4.1 Limitations and Future Directions

Several limitations should be noted. The sample size was modest, and participants displayed heterogeneous brain injury patterns, limiting statistical power and generalizability. Imaging was acquired opportunistically during clinical care, leading to variability in scan quality. Additionally, although advanced modeling approaches improve sensitivity, inherent limitations of diffusion MRI, such as sensitivity to motion and partial volume effects, remain, particularly in unmyelinated neonatal tissue (Dubois et al., 2021). Finally, our cross-sectional design does not allow us to make inferences about the long-term functional implications of observed microstructural differences. Future work should leverage longitudinal follow-up to examine how early microstructural differences predict later neurodevelopmental outcomes. Methodologically, extending these approaches to higher-resolution acquisitions, and combining diffusion metrics with complementary imaging modalities, could further refine the sensitivity and specificity of neonatal microstructural assessment.

### 4.2 Conclusion

Overall, these findings underscore the feasibility and utility of extracting rich microstructural information from routine clinical scans, opening avenues for studying early brain development in preterm infants at scale.

## 5 Statements

### 5.2 Ethics

The studies involving humans were approved by University of Virginia Institutional Review Board for Health Sciences Research. The studies were conducted in accordance with the local legislation and institutional requirements. Written informed consent for participation in this study was provided by the participants’ legal guardians/next of kin.

### 5.3 Author Contributions

BTN: Writing – original draft, Writing – review & editing, Conceptualization, Formal Analysis, Methodology, Visualization; MHP: Writing – original draft, Writing – review & editing, Conceptualization, Formal Analysis, Funding Acquisition, Visualization.

### 5.5 Data availability statement

The raw data supporting the conclusions of this article will be made available by the authors, without undue reservation.

## 5.6 Acknowledgements

We thank Chad Alridge for performing the electronic health record data extraction. ChatGPT (OpenAI, GPT-5.3 model) was used for language editing assistance; all content was reviewed and approved by the authors.

