## Supplement for "A Novel Fixel-Based Approach for Resolving Neonatal White Matter Microstructure from Clinical Diffusion MRI"

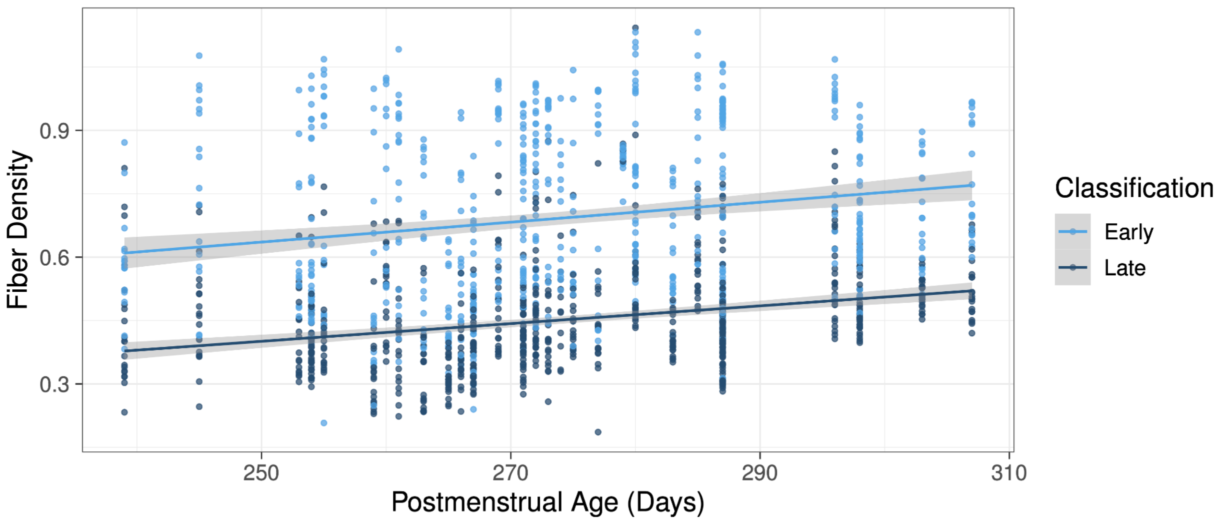


**Figure S1. Association between postmenstrual age and tract class on fiber density in an age-restricted sub-sample.** In a sub-sample of younger infants (postmenstrual age < 360 days), fiber density was higher in early-emerging tracts and increased with postmenstrual age in both early-maturing and later-developing white matter tracts. However, the rate of development of these tracts did not differ in this age-restricted sub-sample, indicating that differences in developmental trajectories between tract classes emerge across a broader developmental window.

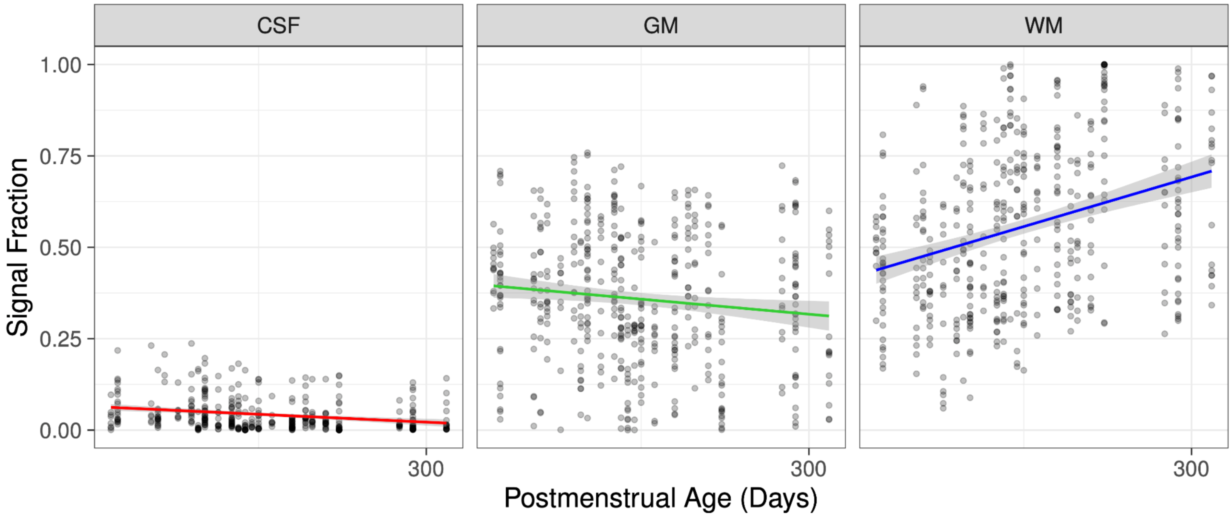


**Figure S2. Associations between postmenstrual age and tissue signal fraction in an age-restricted sub-sample.** In a sub-sample of younger infants (postmenstrual age < 360 days), the significant positive association between postmenstrual age and signal fraction in white matter (WM) remained. However, in this age-restricted sub-sample, the negative association with both grey matter (GM) and cerebrospinal fluid (CSF) were non-significant, indicating that the developmental increase observed in WM signal fraction is robust even within a narrower age range. Red=CSF-like, green=grey matter-like, and blue=white matter-like signal fraction.
